# The Impact of Homologous Recombination Deficiency on First-line Adjuvant Chemotherapy and First-line PARPi Maintenance Therapy in Chinese Patients with Epithelial Ovarian Cancer

**DOI:** 10.1101/2023.01.12.23284477

**Authors:** Lei Li, Yu Gu, Mengpei Zhang, Xiaohua Shi, Zhe Li, Xinyun Xu, Tianqi Sun, Yu Dong, Chao Xue, Xiaoru Zhu, Ran Lv, Kai Jiao, Xuwo Ji, Zhiyong Liang, Ying Jin, Rutie Yin, Ming Wu, Han Liang

**Author notes:** Corresponding authors: H.L., (Lead Contact); M.W.,; R.Y.,; Y.J.,; Z.Liang. These authors contributed equally to this study (Lei Li, Yu Gu, Mengpei Zhang, Xiaohua Shi, Zhe Li).

## Abstract

Homologous recombination deficiency (HRD) testing has been approved by FDA for selecting epithelial ovarian cancer (EOC) patients who may benefit from the first-line poly (ADP-ribose) polymerase inhibitor (PARPi) maintenance therapy. However, the effects of HRD on the clinical outcomes of first-line chemotherapy and first-line PARPi maintenance therapy have not been rigorously evaluated in Chinese EOC patients. Here, we developed an HRD assay and applied it to two large Chinese EOC patient cohorts. In the first-line adjuvant chemotherapy cohort (FACT, N = 380), HRD status significantly improved PFS (median, 15.6 months vs. 9.4 months; HR, 0.688; 95% CI, 0.526 to 0.899; P = 0.003) and OS (median, 89.5 months vs. 60.9 months; HR, 0.636; 95% CI, 0.423 to 0.955; P = 0.008). In the first-line PARPi maintenance therapy cohort (FPMT, N = 83), HRD status significantly improved PFS (median, NA vs 12 months; HR, 0.438; 95% CI, 0.201 to 0.957; P = 0.033) and OS (median, NA vs NA months; HR, 0.12; 95% CI, 0.029 to 0.505; P = 0.001). Our results demonstrate that HRD status is a significant predictor for PFS and OS in both first-line chemotherapy and first-line PARPi maintenance therapy, providing strong real-world evidence for conducting genetic testing and improving clinical recommendations for Chinese EOC patients.

## Introduction

Epithelial ovarian cancer (EOC) is the most lethal gynecologic malignancy (1), and there were more than 313,959 new cases and 207,252 deaths worldwide in 2020 (2). Based on conservative estimates, in 2015, 57,200 new cases and 27,200 deaths occurred in China (3). Identification of mutation carriers among probands not only represents a great opportunity for risk-reduction interventions but also provides reassurance to noncarriers (4). Furthermore, homologous recombination deficiency (HRD) assays have been used to stratify EOC patients for effective treatment (5).

Recent progress in developing targeted therapies of EOC, such as poly (ADP-ribose) polymerase inhibitor (PARPi), has created a great need for genetic testing (6)(7)(8). Four PARPi (i.e., niraparib, olaparib, rucaparib, and fluzoparib) have been approved for the maintenance treatment of patients with EOC who exhibit a complete or partial response to platinum-based chemotherapy regardless of *BRCA* mutation status and HRD status (9)(10)(11)(12)(13)(14)(15)(16). Several HRD models have been developed, including “MyChoice CDx” (17)and “FoundationFocus CDx*BRCA* LOH” (9). However, beyond *BRCA*-mutated tumors, current HRD assays have not demonstrated a differentiation power in predicting patient response to PARPi and other antiangiogenic therapy to justify their routine use in the clinic (5). In particular, despite several recent studies (18)(19)(20)(21)(22), little is known about the real-world impact of HRD on the therapeutic effects of chemotherapy and PARPi in Chinese EOC patients.

To fill this knowledge gap, we developed an HRD assay and applied it to two large patient cohorts to evaluate the impact of HRD in first-line chemotherapy and subsequent first-line PARPi maintenance therapy in Chinese EOC patients.

## Results

### Development and validation of an HRD assay

To characterize the HRD status of tumor samples, we developed a customized sequencing panel named “Precision Human HRD Assay”. This HRD assay contains two sets of probes (**Figure 1A**). One set of HRD-score probes (∼50 K) evenly cover the whole genome (**Figure 1B**) and aims to assess genomic instability on a global scale. We developed an HRD score algorithm to calculate a score for each of the three features: the loss of heterozygosity (LOH), telomeric allelic imbalance (TAI), and large-scale state transitions (LST); and the overall HRD score was the sum of LOH, TAI, and LST scores. The other set of DDR-gene probes aims to evaluate the genotype of 36 DNA damage repair (DDR) genes, among which are 28 homologous recombination repair (HRR) genes (**Table S1**). To validate the analytical performance of our HRD assay, for the same set of samples, we compared our HRD score with that of myChoice-Plus, a widely used commercial HRD assay (R^2^ = 0.983, **Figure 1C**), that based on whole-exome sequencing data (WES) plus backbone (R^2^ = 0.988, **Figure 1D**), and that based on whole-genome sequencing (WGS) data (R^2^ = 0.988, **Figure 1E**), and they all showed extremely high correlations. These results indicate that our HRD assay can quantify the HRD status of tumor samples accurately.

**Figure 1.**
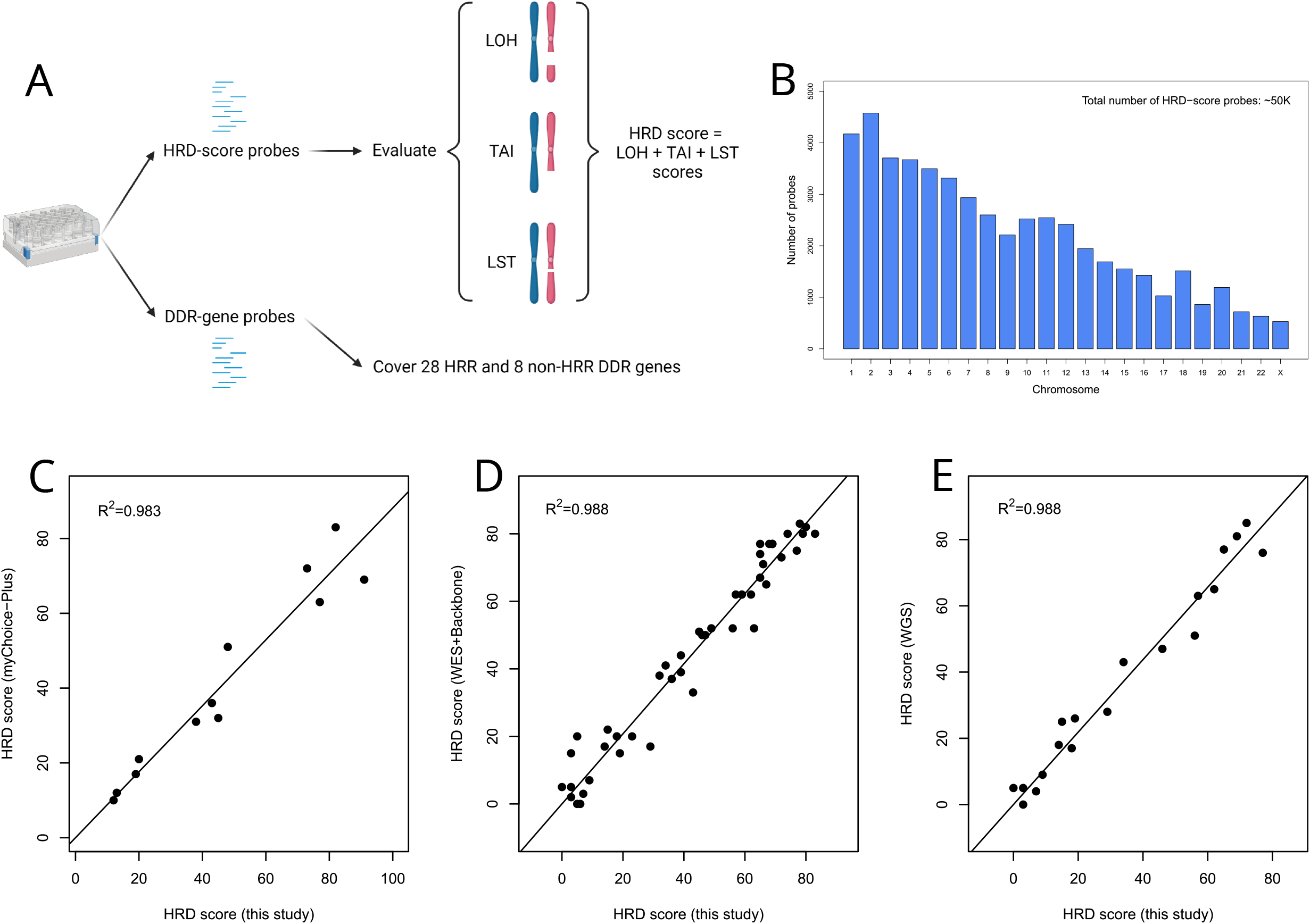
Development and validation of an HRD assay. (A) A diagram showing the components of the Precision Human HRD Assay. (B) Distribution of HRD-score probes by chromosomes. The correlation of the HRD score with those from (C) myChoice-Plus, (D) WES+Backbone, and (E) WGS.

### Tumor characteristics of ECO patients

To evaluate the impact of HRD status on the treatment of Chinese EOC patients, we retrospectively surveyed the medical records of patients from two leading hospitals in China: Peking Union Medical College Hospital (Beijing, China) and West China Hospital of Sichuan University (Chengdu, China). Among the eligible participants who underwent first-line adjuvant chemotherapy, we successfully genotyped 380 patients using our HRD assay and defined them as the FACT cohort (**Table S2**). Among the eligible participants who underwent first-line PARPi maintenance therapy, we successfully genotyped 83 patients and defined them as the FPMT cohort (**Table S3**).

Thus, our study included 463 participants in total (**Figure S1**).

From 36 DDR genes covered by our HRD assay, we further defined several HRR gene subsets, including HRR12, HRR14, HRR26, and HRR28 (**Table S1**). We next examined the mutation status of these genes or subsets in our cohorts. Among all the participants, 172/463 (37.1%) were *BRCA1/2* mutated, and 164/463 (35.4%) had *BRCA1/2* bi-allelic loss-of-function (BILOF). Among 380 participants in the FACT cohort, 133/380 (35%) were *BRCA1/2* mutated, and 130/380 (34.2%) had *BRCA1/2* BILOF. Among 83 participants in the FPMT cohort, 39/83 (47%) were *BRCA1/2* mutated, and 34/83 (41%) had *BRCA1/2* BILOF. In the FACT cohort, 9.2% of the participants were *BRCA1/2* wild-type with at least one of the HRR26 genes mutated; therefore a total of 44.2% participants had at least one of the HRR28 genes mutated, and 55.8% participants had all HRR28 genes as wild-type (**Figure 2A, 2B**). Among all the mutations observed on HRR28 genes, *BRCA1* (58%) and *BRCA2* (13.3%) were the most frequent ones, followed by *RAD51D* (5.9%), *RAD51C* (3.7%), *PTEN* (2.7%), and *PALB2* (2.1%). In the FPMT cohort, we observed a higher frequency of *BRCA1/2* mutations, likely due to the treatment strategy (**Figure 2C, 2D**).

**Figure 2.**
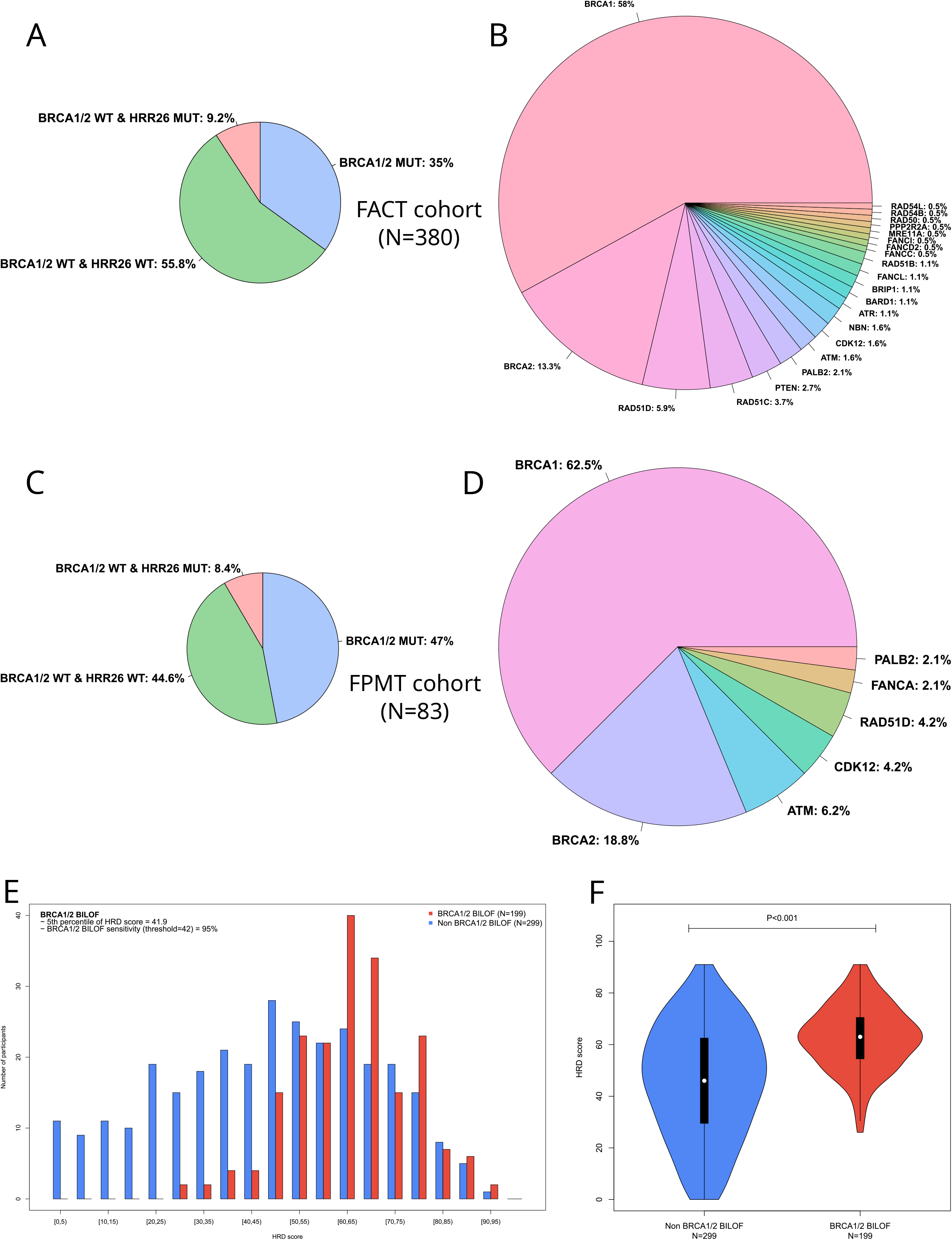
The mutational landscape of homologous recombination repair genes. HRR gene mutation rates in the FACT cohort are summarized (A) by patients and (B) by HRR gene mutations. HRR gene mutation rates in the FPMT cohort are summarized (C) by patients and (D) by HRR gene mutations. Mutation rates in each pie chart sum to 100%. HRR26 is a gene list composed of 26 HRR genes as defined in Table S1. A patient is defined as HRR26-mutated if having at least one of the HRR26 genes mutated. HRD score distribution stratified by *BRCA1/2* BILOF is illustrated (E) by a histogram and (F) by a violin plot. MUT, mutated; WT, wild-type.

Bi-allelic alterations in HRR genes are necessary for loss of function according to the two-hit hypothesis. We tested whether bi-allelic inactivation of HRR genes led to genomic scarring consistent with the underlying DNA-repair deficiency. We observed strong positive associations between BILOF of HRR genes and HRD score for HRR28 (47.7% increase in median HRD score relative to the wild-type; P < 0.001), HRR14 (47.7% increase; P < 0.001), and *BRCA1/2* (44.4% increase; P < 0.001) in the FACT cohort (**Figure S2A**). We further analyzed each individual DDR gene and observed strong positive associations for *BRCA1* (40.8% increase; P < 0.001), *RAD51D* (28.2% increase; P = 0.038), and *TP53* (31.3% increase; P < 0.001) (**Figure S2B**). We finally examined the association between *BRCA1* promoter methylation and HRD score in the FACT cohort and found a positive association (**Figure S3**). We observed a clear distinction in HRD score distribution when stratified by *BRCA1* promoter methylation status (*BRCA1* promoter methylation score ≥ 0.5). These results indicate a good performance of our HRD assay.

To make a binary call on the HRD status of a tumor (i.e., HRD-positive vs. HRD-negative), we aimed to identify a threshold of HRD score that can effectively separate *BRCA1/2* BILOF from the others, as in previous studies (9, 17). The HRD score threshold training set contained 199 BILOF participants in total, including 130 *BRCA1/2* BILOF participants from FACT cohort, 34 *BRCA1/2* BILOF participants from FPMT cohort, and 35 *BRCA1/2* BILOF participants from our routine genetic testing services who met the same eligibility criteria of FACT and FPMT cohort. Within the HRD score threshold training set, the 5th percentile of HRD score was 41.9, for which the sensitivity in predicting *BRCA1/2* BILOF was 95% (**Figure 2E**). Thus, a tumor was defined as HRD-positive if HRD score ≥ 42 or with mutated *BRCA1/2* (**Figure 2F**).

### Effects of HRD status on patient survival time in the FACT cohort

To validate the clinical value of HRD status in EOC patients, we first examined the correlations of HRD status with both chemotherapy-related progression-free survival (PFS) and overall survival (OS) in the FACT cohort. For the PFS analysis stratified by HRD status, the median follow-up time was 35.7 months (IQR 27.3-44.6). Figure 3 shows PFS and OS analyses stratified by *BRCA1/2* mutation and HRD status. The median PFS was 17.7 months for participants with *BRCA1/2* mutation versus 12.1 months for participants with wild-type *BRCA1/2* (HR, 0.629; 95% CI, 0.487 to 0.814; P = 0.003) (**Figure 3A**). When the combination of HRD score and *BRCA1/2* mutation status was used to obtain the HRD status, the median PFS was 15.6 months in HRD-positive participants versus 9.4 months in the HRD-negative participants (HR, 0.688; 95% CI, 0.526 to 0.899; P = 0.003) (**Figure 3B**). Even in the *BRCA1/2* wild-type subgroup, there was an improved PFS in participants with a positive HRD status (median, 13 months versus 9.4 months; HR, 0.769; 95% CI, 0.578 to 1.023; P = 0.071) (**Figure 3C**). We observed similar improvements in OS for participants who were *BRCA1/2* mutated, HRD-positive, and HRD-positive with wild-type *BRCA1/2*. The magnitude of the OS benefit was most prominent in *BRCA1/2* mutated participants (median, 90.9 months versus 70.5 months; HR, 0.6; 95% CI, 0.39 to 0.922; P = 0.004) (**Figure 3D**), followed by the HRD-positive (median, 89.5 months versus 60.9 months; HR, 0.636; 95% CI, 0.423 to 0.955; P = 0.008) (**Figure 3E**), and the least in *BRCA1/2* wild-type participants with a positive HRD status (median, 74.9 months versus 60.9 months; HR, 0.724; 95% CI, 0.468 to 1.122; P = 0.147) (**Figure 3F**).

**Figure 3.**
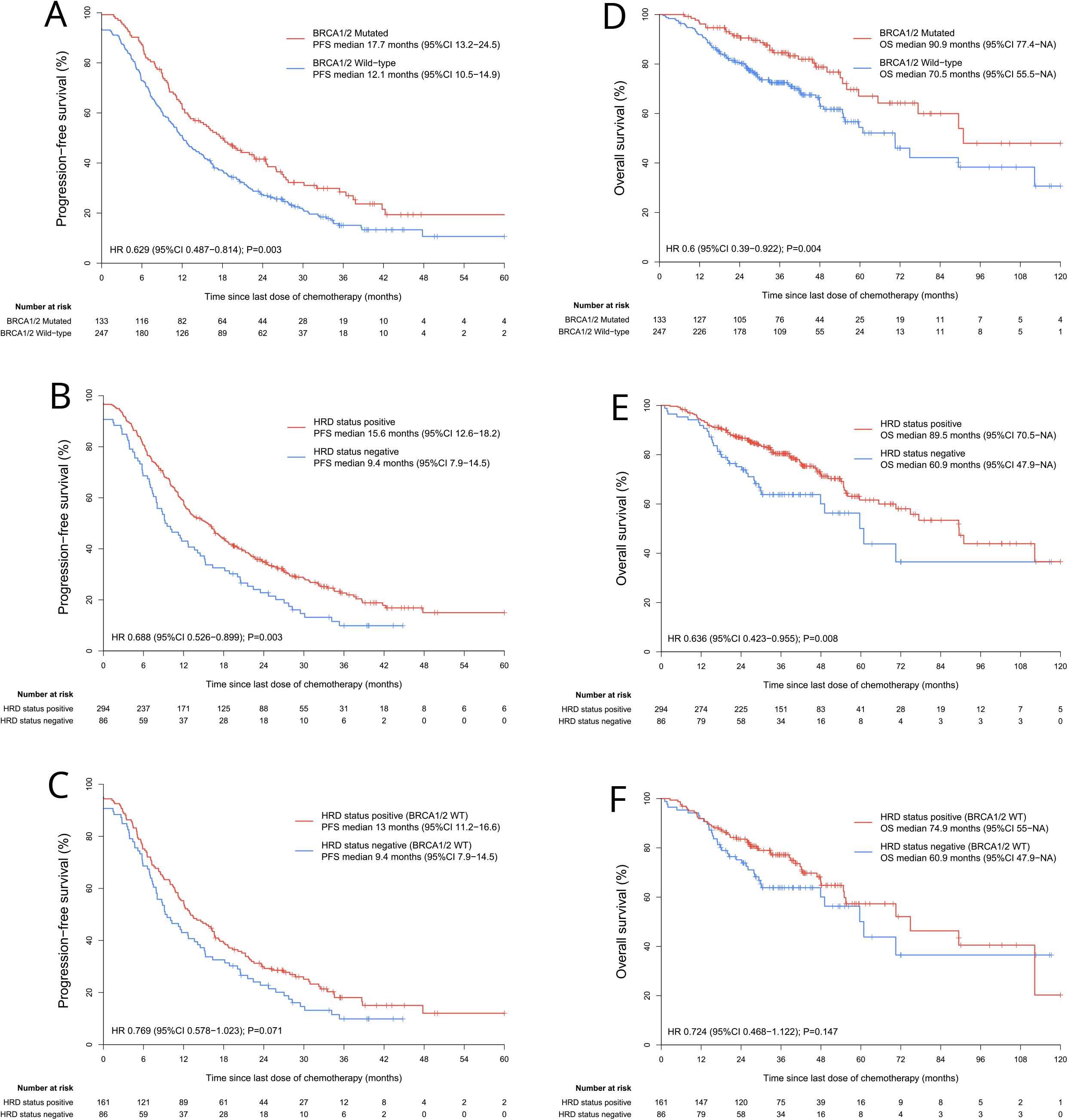
FACT cohort patient survival analyses stratified by HRD status. PFS analyses in the FACT cohort stratified by (A) *BRCA1/2* mutation, (B) HRD status, and stratified by (C) HRD status in *BRCA1/2* wild-type participants are presented. OS analyses in the FACT cohort stratified by (D) *BRCA1/2* mutation, (E) HRD status, and stratified by (F) HRD status in *BRCA1/2* wild-type participants are presented. WT, wild-type.

We next performed PFS and OS analyses on HRD-positive participants stratified by different causes of HRD. Participants whose tumors were HRD-positive due to genetic changes (*BRCA1/2* mutation) outperformed the other two groups in both PFS and OS (**Figure S4**). Participants whose tumors were HRD-positive due to epigenetic changes (*BRCA1/2* wild-type and *BRCA1* promoter methylation status high) showed similar PFS and OS compared with those whose tumors were HRD-positive due to unknown reasons (*BRCA1/2* wild-type, *BRCA1* promoter methylation status low, and HRD status positive).

Finally, we investigated whether HRD status could predict the platinum sensitivity status in the FACT cohort. *BRCA1/2* mutations were strongly associated with being platinum-sensitive: 87.2% of participants with *BRCA1/2* mutations were platinum-sensitive, whereas only 72.9% of participants without *BRCA1/2* mutations were platinum-sensitive (P = 0.001) (**Figure S5A**). HRD status also increased the platinum sensitivity rate (80.6% versus 68.6%, P = 0.026) (**Figure S5B**). HRD status achieved a smaller increase in platinum sensitivity rate in the *BRCA1/2* wild-type participants (75.2% versus 68.6%, P = 0.295) (**Figure S5C**).

### Effects of HRD status on patient survival time in the FPMT cohort

To further assess whether HRD status could stratify Chinese EOC patients for PARPi maintenance therapy in the first-line setting, we analyzed PFS and OS in FPMT stratified by *BRCA1/2* mutation and HRD status (**Figure 4**). For the PFS analysis stratified by HRD status, the median follow-up time was 22.1 months (IQR 10.9-29.3). *BRCA1/2* mutation was a strong predictor for PFS (median, NA versus 20 months; HR, 0.423; 95% CI, 0.198 to 0.906; P = 0.023) (**Figure 4A**), but not as much for OS (median, NA versus NA; HR, 0.592; 95% CI, 0.141 to 2.477; P = 0.467) (**Figure 4D**). There was a significantly improved PFS in HRD-positive participants (median, NA versus 12 months; HR, 0.438; 95% CI, 0.201 to 0.957; P = 0.033) (**Figure 4B**), and HRD status also showed an obvious benefit in OS (median, NA versus NA; HR, 0.12; 95% CI, 0.029 to 0.505; P = 0.001) (**Figure 4E**). Furthermore, in the *BRCA1/2* wild-type subgroup, HRD status also demonstrated a significant benefit in OS (**Figure 4F**).

**Figure 4.**
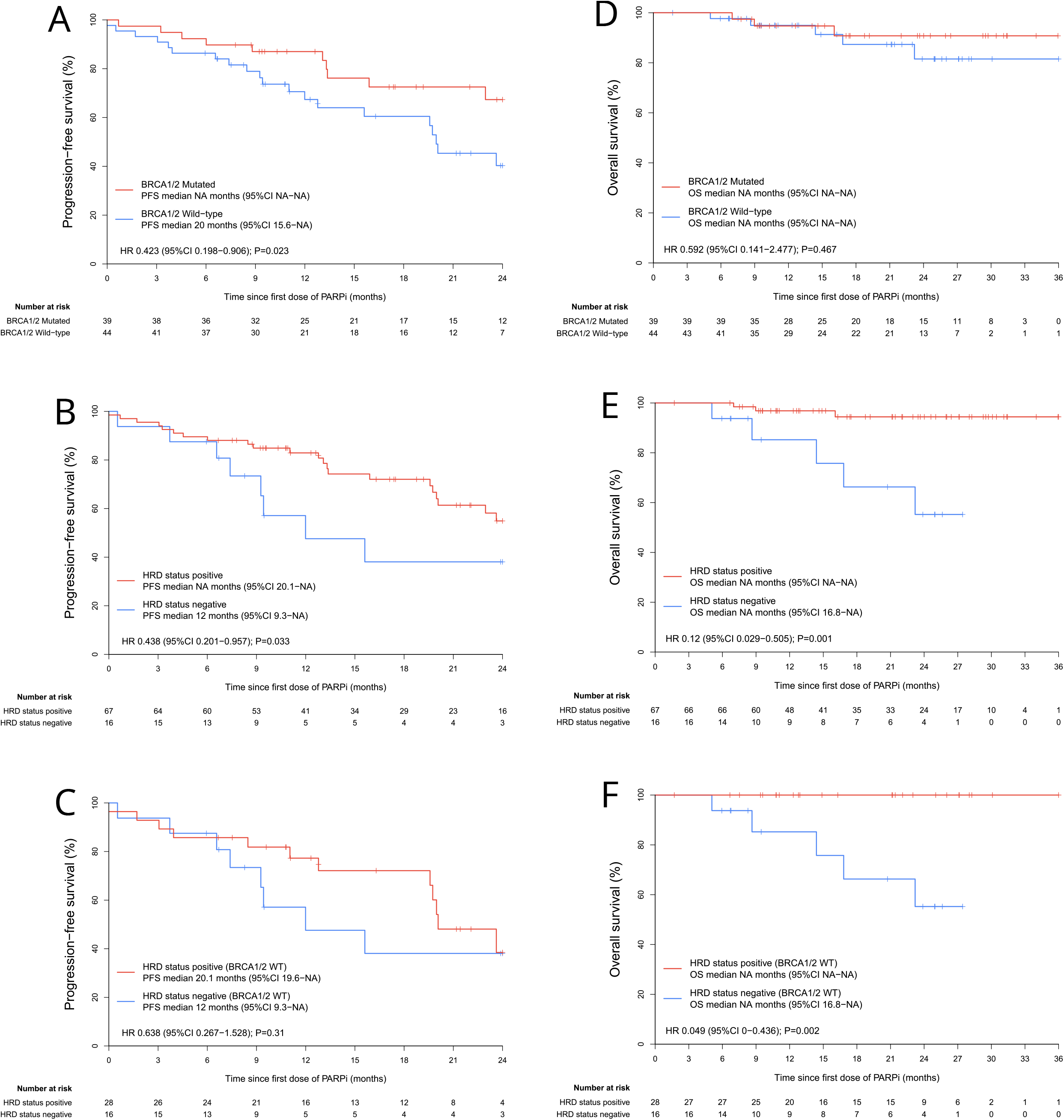
FPMT cohort patient survival analyses stratified by HRD status. PFS analyses in the FPMT cohort stratified by (A) *BRCA1/2* mutation, (B) HRD status, and stratified by (C) HRD status in *BRCA1/2* wild-type participants are presented. OS analyses in the FPMT cohort stratified by (D) *BRCA1/2* mutation, (E) HRD status, and stratified by (F) HRD status in *BRCA1/2* wild-type participants are presented. WT, wild-type.

### HRD status versus HRR genes in *BRCA1/2* wild-type participants

We investigated potential predictors for efficacy in *BRCA1/2* wild-type participants (**Figure 5**). For the PFS analyses in the FACT cohort, HRD status, HRR28, and HRR14 were all satisfactory predictors (**Figure 5A**). For OS analyses in the FACT cohort, HRR28 and HRR14 performed slightly worse compared to HRD status (**Figure 5B**). For both PFS and OS analyses in FPMT cohort, HRR14 and HRR28 were poor predictors (**Figure 5C, 5D**). In the FPMT cohort, HRD status was a strong predictor for both PFS and OS (**Figure 5C, 5D**). Collectively, these results suggest that HRD status was a better predictor for efficacy than the mutational status of HRR genes in *BRCA1/2* wild-type participants.

**Figure 5.**
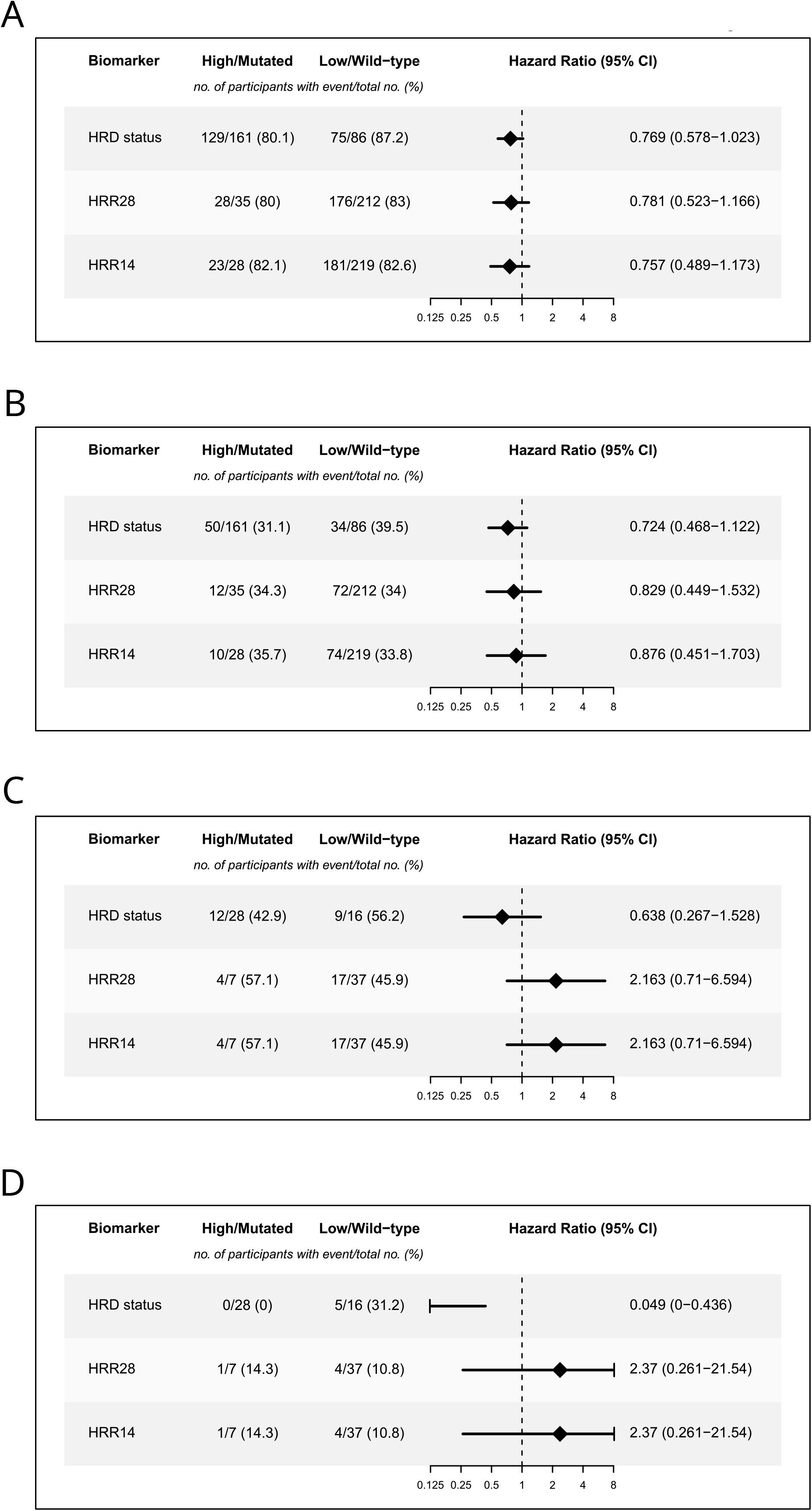
Predictors for efficacy in *BRCA1/2* wild-type participants. Forest plots illustrate whether HRD status, HRR28, and HRR14 were strong predictors for efficacy in *BRCA1/2* wild-type participants. Explorations were split by (A) PFS in the FACT cohort, (B) OS in the FACT cohort, (C) PFS in the FPMT cohort, and (D) OS in the FPMT cohort.

We further explored whether alterations of each gene were associated with HRD status in *BRCA1/2* wild-type participants. In the combined dataset (FACT and FPMT cohorts combined), *TP53* BILOF was significantly enriched in HRD-positive participants in the *BRCA1/2* wild-type subgroup. In contrast, the frequency of *TP53* mono-allelic loss-of-function (MOLOF) was higher in the HRD-negative group, but when considering the co-occurrence of *BRCA1*-LOH, the trend was reversed (**Figure S6**). *TP53* BILOF did not show any predictive value for efficacy in FACT or FPMT cohort (**Figure S7**).

## Discussion

Here we developed a rigorous HRD assay and assessed the impact of HRD status on the efficacy of first-line chemotherapy and PARPi maintenance therapy in Chinese patients with primary EOC. Our HRD scoring system demonstrates its utility in predicting therapy efficacy in Chinese EOC patients. Furthermore, consistent with the results in the PRIMA trial (8) and ATHENA-MONO (23), even in *BRCA* wild-type patients, HRD status in our study demonstrated significant survival benefits in both PFS and OS when appropriate HRD score threshold was selected. Despite some fundamental disparities between the FACT and FPMT cohorts, including different mechanisms of drug resistance and patient pharmacogenomic features (e.g., intratumor heterogeneity and HRR gene mutation rate) (24)(25)(26)(27)(28), our HRD assay demonstrates a robust predictive power.

Our study shows a moderately high percentage of being HRD-positive in the *BRCA1/2* wild-type participants, which is similar to the findings from Japan (29)(30) and another study in China (31) but higher than those reported in previous studies (8)(17). Sinha et al. (32) demonstrated higher genomic instability, HRD and chromothripsis among tumors from African Americans across many cancer types (including ovarian cancer) compared with European Americans in The Cancer Genome Atlas. Hsiao et al. (33) reported that genome-wide HRD scores showed racial differences between Caucasian population and Asian population across cancer types, with a significantly higher score observed in Asian population. Furthermore, *BRCA1/2* mutation rates reached 35% in our study, which was slightly higher than other reports from China (34)(35) and higher than other countries (36)(37). Ethnic-specific *BRCA1/2* variation in Asian populations may be explained as follows (38): low overlapping between Asian and non-Asian *BRCA1/2* variants, low overlapping within different Asian populations, different variation spectrum and variation types, and differences in *BRCA1/2* founder mutations. *BRCA1* c.5470_5477del, which is a reported Chinese *BRCA1* founder mutation (39), has high occurrence in patients from our routine genetic testing services and from patients in this study. Taken together, our results suggest that the genetic ancestry origin is a major factor for the higher rate of HRD positive observed in this study.

Consistent with a previous report (30), we show that among the HRD-positive participants, HRD status positive caused by *BRCA1/2* mutations and *BRCA1/2* BILOF (data not shown) demonstrated a better prognosis than those caused by epigenetic changes or unknown reasons. These findings suggest that *BRCA1/2* alterations contributed greatly to the HRD status. Contrary to *BRCA1/2* mutations, mutations in other HRR genes had no inherent association with survival outcomes in the FACT cohort. These findings are compatible with previous studies and current recommendations (5)(40)(41). We found that higher *BRCA1* methylation score was associated with higher HRD scores. However, in this and previous studies (30), epigenetic changes in HRR genes were less associated with survival outcomes compared with genetic HRD. Furthermore, we found that *TP53-*mutated participants had higher median HRD scores than the *TP53* wild-type, especially for *TP53* BILOF, which was significantly enriched in high HRD score participants in the *BRCA1/2* wild-type subgroup. These findings are consistent with previous observations in prostate cancer (42) and pan-cancer analysis (43). Loss of *TP53* may lead to chromosomal instability and aneuploidy (44), which is associated with a higher HRD score.

In conclusion, we developed an effective HRD assay for Chinese EOC patients, which provides precise predictions for first-line chemotherapy and PARPi efficacy, including PFS and OS. The HRD status shows high accordance with *BRCA1/2* mutations, BILOF of HRR genes, and *BRCA1* promoter methylation scores. These findings should be further validated in future cohorts or RCT trials with larger sample sizes and universal treatment protocols.

## Methods

### HRD assay development

The Precision Human HRD Assay contains two sets of probes, HRD-score probes (∼50K) and DDR-gene probes, which were used to evaluate HRD score and genotype 36 DDR genes (**Table S1**), respectively. To validate the analytical performance, HRD score was compared with the scores of myChoice-Plus, WES+Backbone, and WGS. The WES+Backbone (IDTxGen Exome Research Panel v1.0, IDT xGen Human CNV Backbone Hybridization Panel) and WGS data were sequenced on NovaSeq platform in PE150 mode by mean depth of 150× (tumor) and 50× (control). The WES+Backbone and WGS data was analyzed using FACETS (45) and scarHRD (46).

The SNPs targeted by the Precision Human HRD Assay were selected based on the following criteria: SNPs on Y chromosome were removed; mitochondrial SNPs were removed; SNPs with minor-allele-frequency (MAF) less than 1% in European or West African population were removed; SNPs with MAF less than 5% in East Asian population were removed; SNPs significantly deviated from Hardy-Weinberg Equilibrium (HWE) in any of the three populations mentioned above were removed; SNPs with Fst (fixation indices) < 0.05 in East Asian population were removed; SNPs with CG-content < 40% or >60% were removed; SNPs located on Short Tandem Repeats (STRs) were removed; SNPs evenly covered the human genome.

We developed in-house HRD score algorithm to assess genomic instability and calculated a score for each of the three features: LOH (47), TAI (48), and LST (49). HRD score was the sum of LOH, TAI, and LST scores. Sequencing data were aligned to the human reference genome (GRCh37/UCSC hg19), and a custom bioinformatic analysis pipeline was used to detect single nucleotide variants (SNVs) and small insertions and deletion (indels) mutations in protein-coding regions and intron/exon boundaries of the 36 genes. Variants were classified according to the recommendations of the American College of Medical Genetics and Genomics (ACMG) for standards in the interpretation of sequence variations. Clinically significant variants were classified as “class 5: pathogenic” or “class 4: likely pathogenic.”

### EOC participants

The study was approved by the Ethics Committee of Peking Union Medical College Hospital, Beijing (project ID: JS–1932). Medical records of EOC patients from Peking Union Medical College Hospital and West China Hospital of Sichuan University were surveyed retrospectively. Eligibility criteria included being at least 18 years old at diagnosis; a histological/pathologic diagnosis of EOC (ovarian cancer, fallopian tube cancer, or primary peritoneal cancer); high-grade serous or grade-3 endometroid histological subtype; FIGO stage II, III, or IV; first-line surgery performed; administered at least 5 rounds of first-line chemotherapy. Eligibility criteria unique to the FACT cohort included no maintenance therapy of any kind; no PARPi administered during first-line therapies for any purpose; and the date of the last dose of first-line adjuvant chemotherapy between 2009/12/01 and 2020/05/01. The FACT cohort patients were collected from two different investigators. Eligibility criteria unique to FPMT included achieving complete or partial response after first-line adjuvant chemotherapy and PARPi administered as first-line maintenance therapy. FPMT cohort patients were collected from two different investigators.

### Molecular analyses of patient samples

For each participant, at least 10 slides of 5μm formalin-fixed paraffin-embedded (FFPE) tumor samples were collected with the patient’s informed consent. The first slide was stained with hematoxylin and eosin (H&E). The H&E slide was reviewed by two independent pathologists to determine the histological type and neoplastic cellularity (30% minimum). Genomic deoxyribonucleic acid (gDNA) was extracted and quantified from the patient’s specimen(s) using standardized methodology. Tumor-only genetic testing was conducted with the Precision Human HRD Assay. Among the 36 DDR genes, we further defined several HRR gene lists (**Table S1**). If at least one gene in a gene list was mutated (class 5 or 4), then the gene list was considered mutated. An additional 100ng genomic DNA was used to evaluate *BRCA1* promoter methylation status via bisulfite sequencing. We defined the average methylation ratio across CpG sites (i.e., *BRCA1* promoter methylation score) ≥ 0.5 as high methylation status.

### Endpoints

For the FACT cohort, the primary endpoint was PFS, defined as the time from the last dose of chemotherapy to disease progression or death, whichever occurred first. Participants with uncontrolled disease, defined as the participants whose disease progressed during first-line adjuvant chemotherapy, had PFS time set as zero and censoring status set as event occurred. Secondary endpoints included OS, defined as the time from the last dose of chemotherapy to death, and platinum sensitivity status (PSS), defined as whether progression-free survival was longer than 6 months. For the FPMT cohort, the primary endpoint was PFS, defined as the time from the first dose of PARPi to disease progression or death, whichever occurred first. The secondary endpoint was OS, defined as the time from the first dose of PARPi to death. The cutoff date for assessing disease progression and survival of participants was February 23, 2022.

### HRD score threshold training

The HRD score threshold training set was composed of three parts: *BRCA1/2* BILOF participants from FACT cohort, *BRCA1/2* BILOF participants from FPMT cohort, and selective *BRCA1/2* BILOF participants from our routine genetic testing services who met the shared eligibility criteria of FACT and FPMT cohort. The third part of HRD score threshold training set was added to further expand the number of *BRCA1/2* BILOF participants, since larger training set brings more robust HRD score threshold. The HRD score threshold was trained by reaching 95% sensitivity in predicting *BRCA1/2* BILOF, i.e., the HRD score threshold was the 5th percentile of HRD score in *BRCA1/2* BILOF participants. *BRCA1/2* BILOF was defined as meeting one of the following three criteria: (i) one allele as class 4/5 mutated, and the other allele as LOH, (ii) two class 4/5 mutations in the same gene, or (iii) one allele as methylation status high and the other allele as LOH.

### Statistical analyses

Statistical analyses were conducted using R 4.2.1. PFS and OS were estimated using the Kaplan-Meier method. Treatment effect differences were assessed using the log-rank test. In the FACT cohort, HR and associated 95% CI were calculated using the CoxPH model, adjusted for age, FIGO stage, surgery residual, surgery type, pre-treatment CA125, concurrent use of bevacizumab, and the round of chemotherapy. For analyses in the FPMT cohort and subgroup analyses in the FACT cohort, HR and associated 95% CI were calculated using the unadjusted CoxPH model. If one arm in PFS or OS analysis had zero events, Firth’s penalized maximum likelihood bias reduction method for the CoxPH model was applied. Fisher’s exact test was used to assess whether the proportion of platinum-sensitive participants was significantly different between strata.

### Data Availability

Raw sequencing data files of patients cannot be publicly shared under the obtained institutional review board approval, as patients were not consented to share raw sequencing data beyond the research and clinical terms. However, the datasets generated and analyzed during the current study are available from the corresponding author on reasonable request and upon a data usage agreement.

## Data Availability

All data produced in the present study are available upon reasonable request to the authors

## Acknowledgments

This study was supported by a grant from CAMS Innovation Fund for Medical Sciences (CIFMS-2017-I2M-1-002). We thank K. Mojumdar for editorial assistance.

## Author Contributions

X.J., Z. Liang, Y.J., R.Y., M.W. and H.L. conceived and designed the study; L.L., Y.G., M.Z., X.S., Y.D., K.J., Z. Liang, Y.J., R.Y. and M.W. contributed to sample collection and result interpretation, Z. Li, X.X., T.S. and Y.D. performed data analysis; C.X., X.Z. and R.L. performed experiments; L.L., Z. Li, X.X., T.S. and H.L. wrote the manuscript with inputs from other authors.

