## Supplemental Figures and Tables for "The Impact of Homologous Recombination Deficiency on First-line Adjuvant Chemotherapy and First-line PARPi Maintenance Therapy in Chinese Patients with Epithelial Ovarian Cancer"

### Supplemental Information

#### Supplemental Figures

- Figure S1. The overview of patient cohorts included in this study
- Figure S2. Association between HRR gene alterations and HRD score in FACT cohort
- Figure S3. Association between *BRCA1* promoter methylation and HRD score in FACT cohort
- Figure S4. The patient survival analysis stratified by different causes of HRD in FACT cohort
- Figure S5. The platinum sensitivity status stratified by HRD status in FACT cohort
- Figure S6. Association between TP53 alteration and HRD status in *BRCA1/2* wild-type participants
- Figure S7. The patient survival analysis stratified by TP53 BILOF

#### Supplemental Tables

- Supplemental Table S1. DDR genes and HRR gene lists
- Supplemental Table S2. Baseline clinical characteristics in FACT cohort
- Supplemental Table S3. Baseline clinical characteristics in FPMT cohort

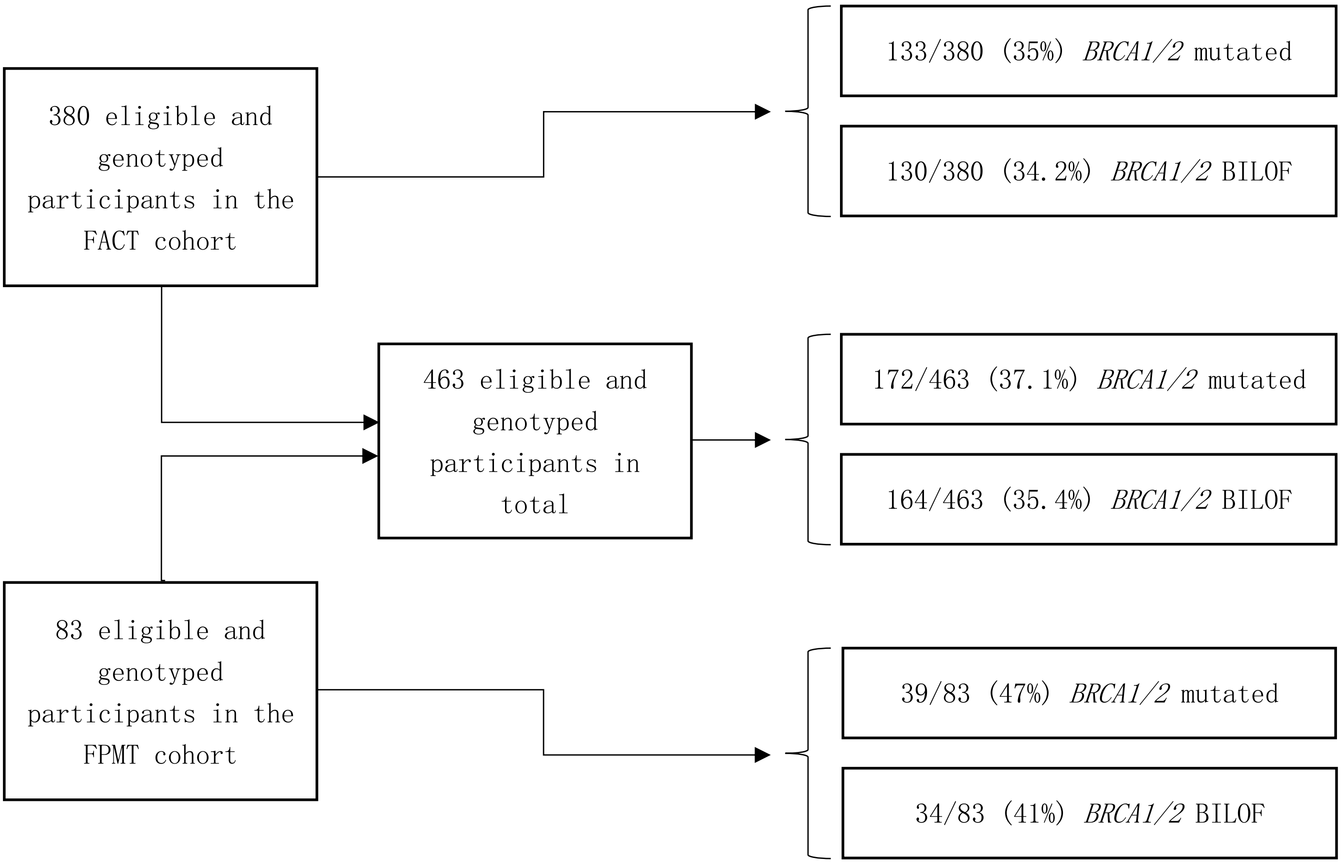


**Figure S1. The overview of patient cohorts included in this study**

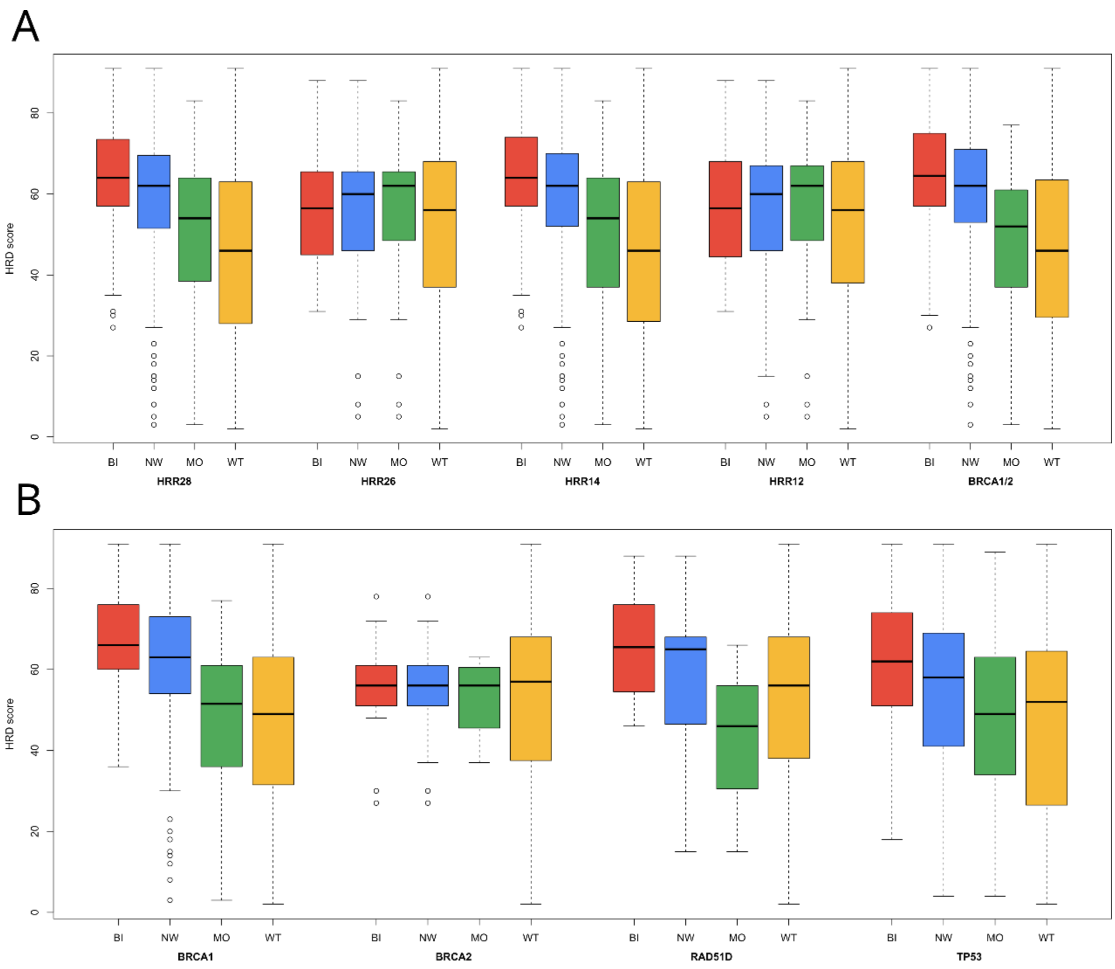


**Figure S2. Association between HRR gene alterations and HRD score in FACT cohort**

(A, B) Boxplots illustrate the distribution of HRD score stratified by alteration type of (A) HRR gene lists, and (B) selective DDR genes. Alteration type includes BI, MO, NW, and WT. BI, bi-allelic loss-of-function; MO, mono-allelic loss-of-function; NW, non-wild-type; and WT, wild-type.

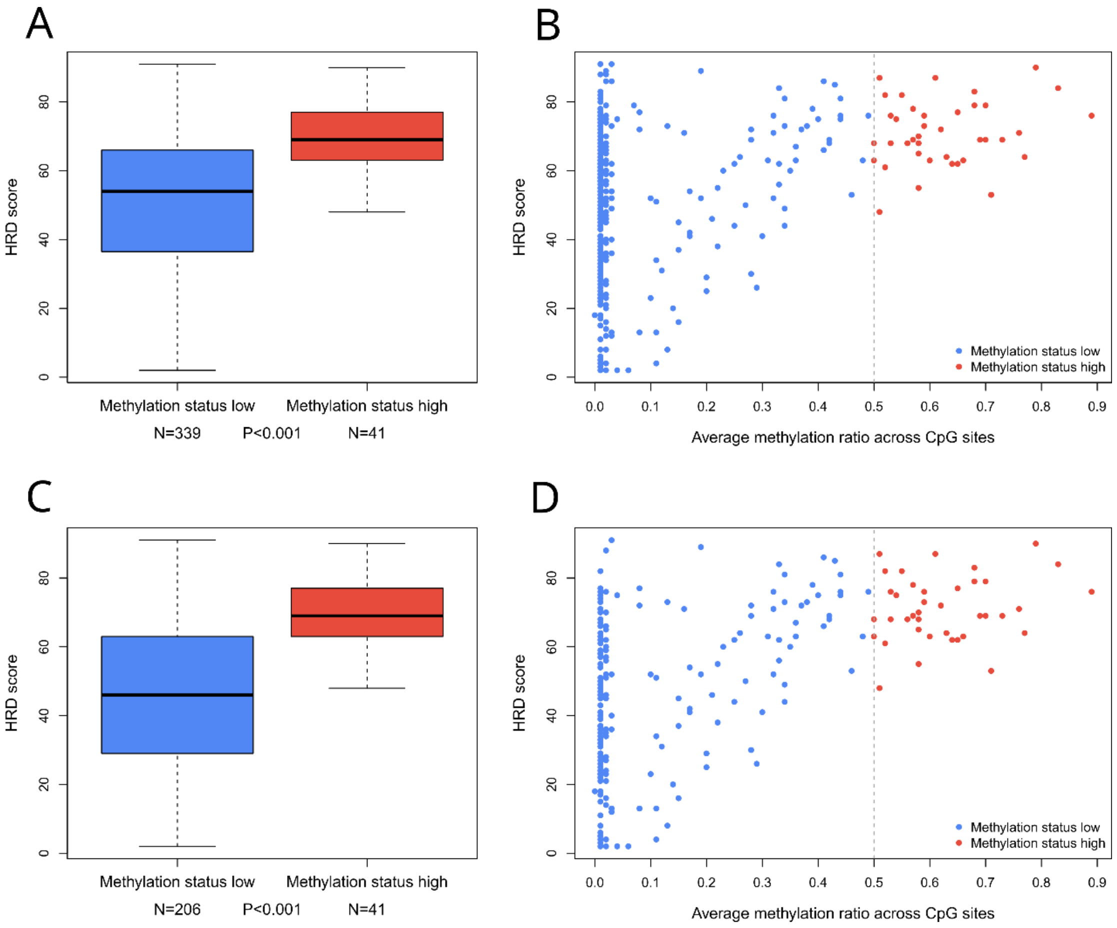


**Figure S3. Association between *BRCA1* promoter methylation and HRD score in FACT cohort**

Boxplots illustrate the distribution of HRD score stratified by *BRCA1* promoter methylation status (*BRCA1* promoter methylation score ≥ 0.5 or not) in (A) FACT cohort and (C) *BRCA1/2* wild-type participants in the FACT cohort. Scatter plots illustrate the relationship between *BRCA1* promoter methylation score and HRD score in (B) FACT cohort and (D) *BRCA1/2* wild-type participants in the FACT cohort. P-values were calculated by Wilcoxon test.

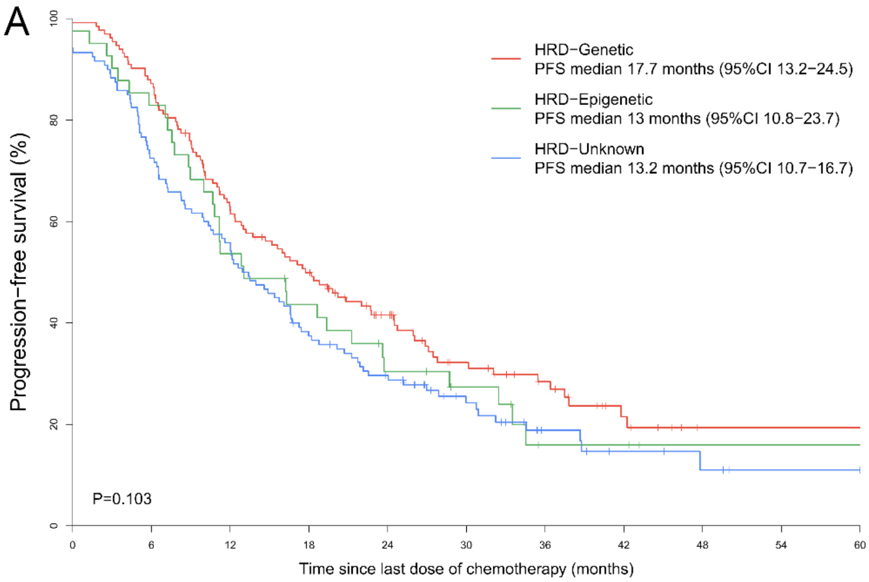


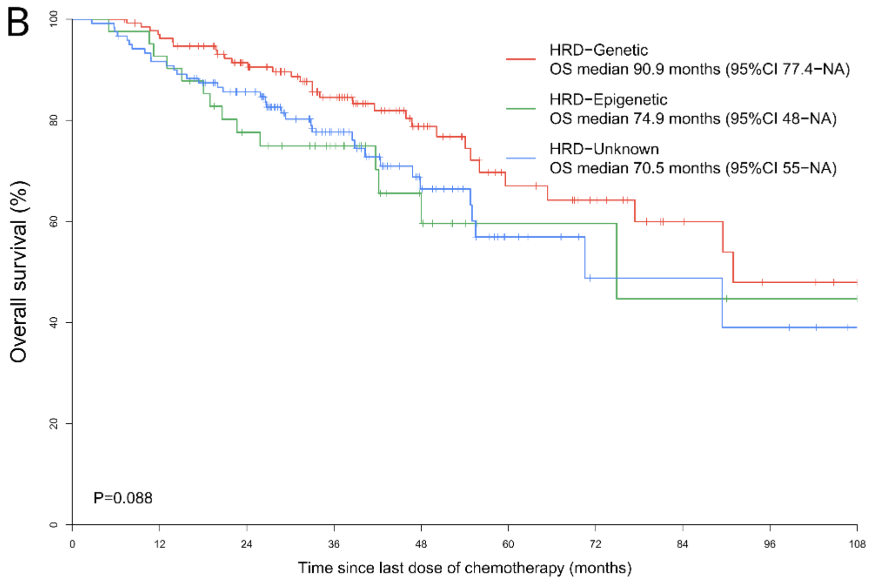


**Figure S4. The patient survival analysis stratified by different causes of HRD in FACT cohort**

(A) PFS and (B) OS analyses stratified by different causes of HRD in HRD-positive participants in the FACT cohort. HRD-Genetic represents HRD-positive participants caused by genetic changes (*BRCA1/2* mutated); HRD-Epigenetic represents HRD-positive participants caused by epigenetic changes (*BRCA1/2* wild-type, and *BRCA1* promoter methylation status high); HRD-Unknown represents HRD-positive participants caused by unknown reasons (*BRCA1/2* wild-type, *BRCA1* promoter methylation status low, and HRD status positive). P-values were calculated by log-rank test.

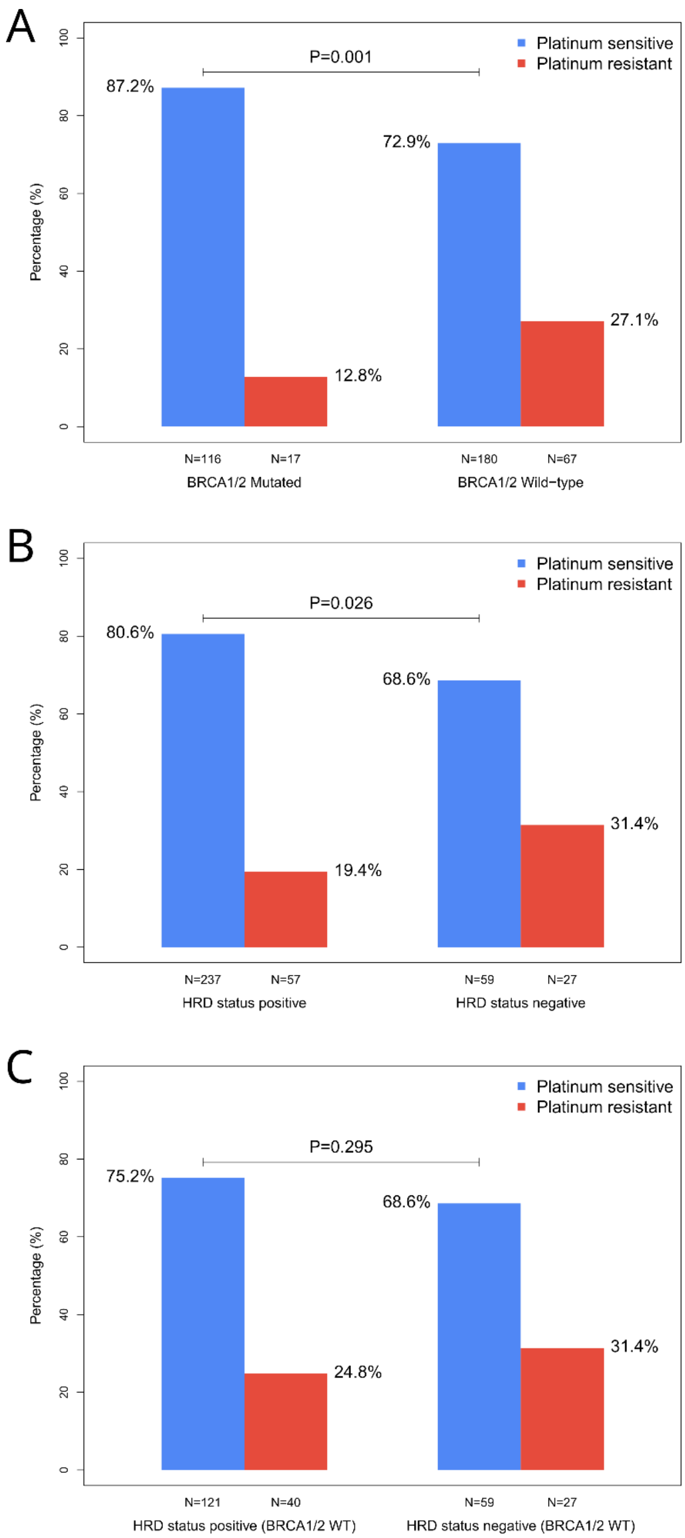


**Figure S5. The platinum sensitivity status stratified by HRD status in FACT cohort** (A-C) PSS analyses in FACT cohort stratified by (A) *BRCA1/2* mutation, (B) HRD status, and (C) HRD status in *BRCA1/2* wild-type participants. WT, wild-type.

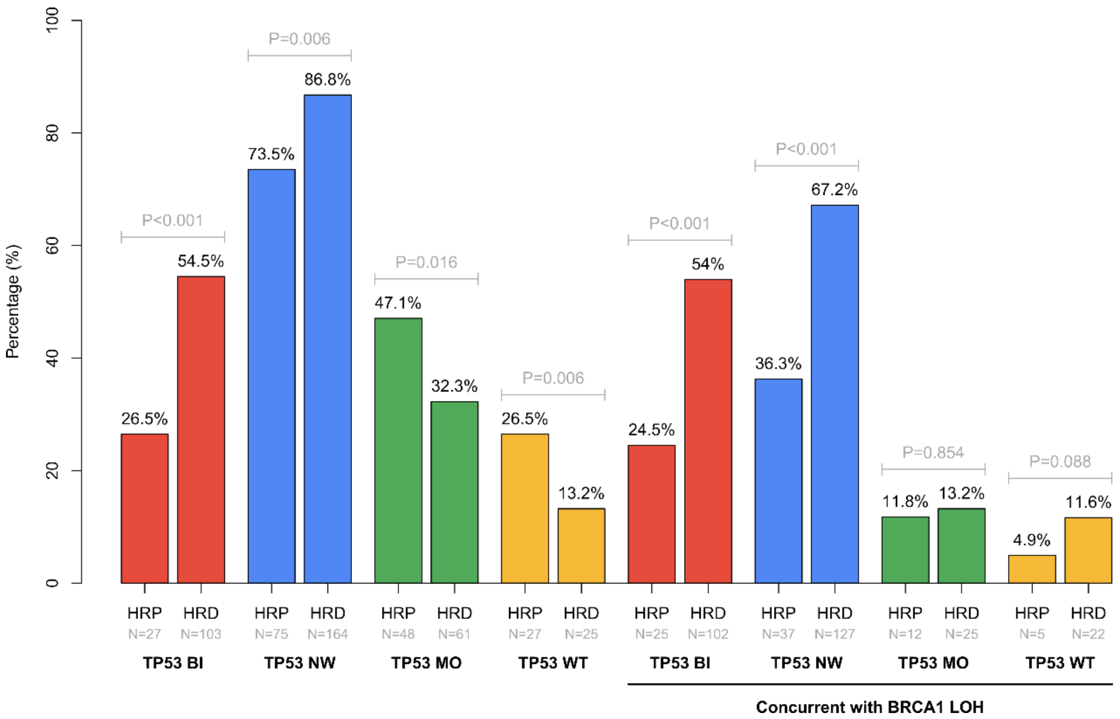


**Figure S6. Association between TP53 alteration and HRD status in *BRCA1/2* wild-type participants**

Barplots illustrate the percentage of HRP (HRD-negative) and HRD (HRD-positive) participants under different alteration types of *TP53*, and under different alteration types of *TP53* concurrent with *BRCA1* LOH. Alteration type includes BI, MO, NW, and WT. P-values were calculated by Fisher's exact test. BI, bi-allelic loss-of-function; MO, mono-allelic loss-of-function; NW, non-wild-type; WT, wild-type; HRP, homologous recombination proficient (HRD-negative); HRD, homologous recombination deficient (HRD-positive); LOH, loss-of-heterozygosity.

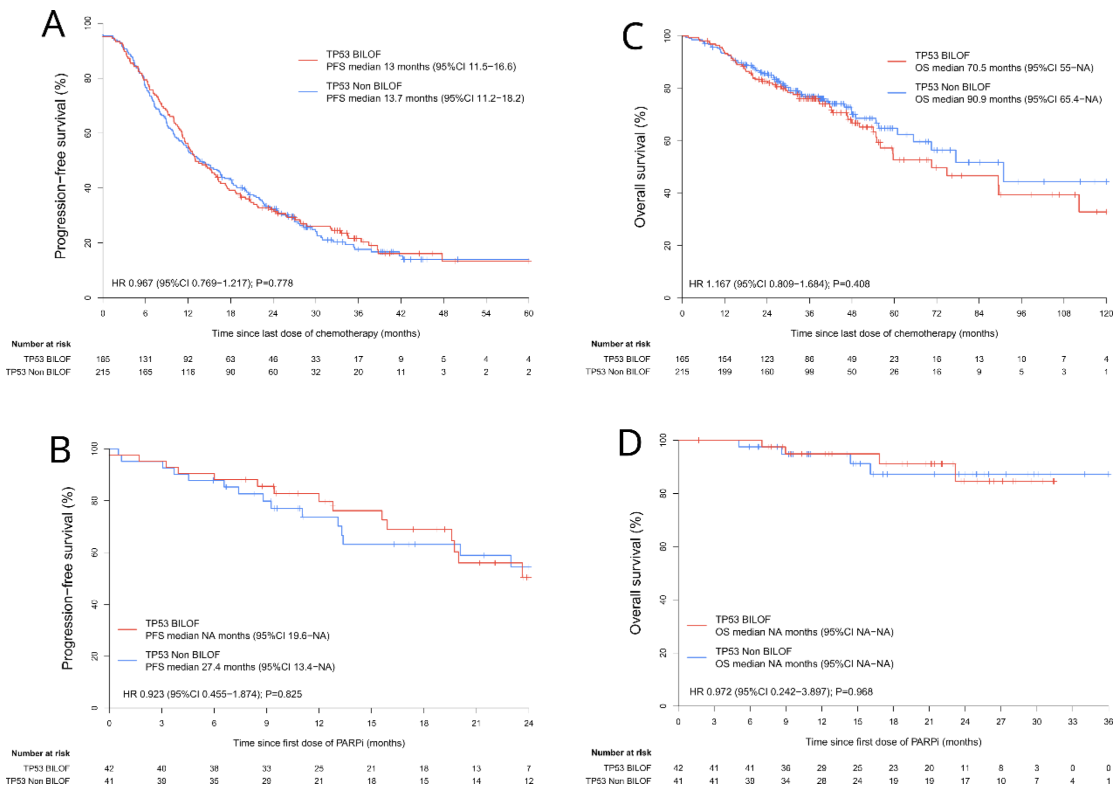


**Figure S7. The patient survival analysis stratified by TP53 BILOF**

(A, B) PFS analyses stratified by *TP53* BILOF in (A) FACT cohort and (B) FPMT cohort. (C, D) OS analyses stratified by *TP53* BILOF in (C) FACT cohort and (D) FPMT cohort.

### Table S1. DDR* genes and HRR gene lists

| DDR* genes | HRR gene lists | | | |
|---|---|---|---|---|
| | HRR28 | HRR26 | HRR14 | HRR12 |
| *ATM* | √ | √ | √ | √ |
| *ATR* | √ | √ | | |
| *BAP1* | √ | √ | | |
| *BARD1* | √ | √ | √ | √ |
| *BRCA1* | √ | | √ | |
| *BRCA2* | √ | | √ | |
| *BRIP1* | √ | √ | √ | √ |
| *CDH1* | | | | |
| *CDK12* | √ | √ | √ | √ |
| *CHEK1* | √ | √ | √ | √ |
| *CHEK2* | √ | √ | √ | √ |
| *EMSY*** | √ | √ | | |
| *EPCAM* | | | | |
| *FAM175A* | √ | √ | | |
| *FANCA* | √ | √ | | |
| *FANCC* | √ | √ | | |
| *FANCD2* | √ | √ | | |
| *FANCI* | √ | √ | | |
| *FANCL* | √ | √ | √ | √ |
| *MLH1* | | | | |
| *MRE11A* | √ | √ | | |
| *MSH2* | | | | |
| *MSH6* | | | | |
| *NBN* | √ | √ | | |
| *PALB2* | √ | √ | √ | √ |
| *PMS2* | | | | |
| *PPP2R2A* | √ | √ | | |
| *PTEN* | √ | √ | | |
| *RAD50* | √ | √ | | |
| *RAD51B* | √ | √ | √ | √ |
| *RAD51C* | √ | √ | √ | √ |
| *RAD51D* | √ | √ | √ | √ |
| *RAD54B* | √ | √ | | |
| *RAD54L* | √ | √ | √ | √ |
| *STK11* | | | | |
| *TP53* | | | | |

Note: If at least one gene in a gene list is mutated, then the gene list is defined as being mutated.

*Most genes listed above are involved in HRR and mismatch repair pathway, except for *STK11*, *TP53*, *CDH1*.

**Amplification and overexpression of *EMSY*, as an alternative way by which tumors selectively inactivate the BRCA pathway (Bell et al., 2011), indicate its mechanism for HRD is different from the “loss of function” of other HRR genes. However, *EMSY* amplification analysis was not performed in this study.

### Table S2. Baseline clinical characteristics in FACT cohort

| Characteristic | HRD status negative (N=86) | HRD status positive (N=294) |
|---|---|---|
| Age, years | 54 (48-62) | 53 (47-61) |
| Cancer type | | |
| Fallopian tube cancer | 8 (9.3) | 34 (11.6) |
| Ovarian cancer | 78 (90.7) | 258 (87.8) |
| Primary peritoneal cancer | 0 (0) | 2 (0.7) |
| Histological type | | |
| Grade-3 endometroid | 3 (3.5) | 5 (1.7) |
| High-grade serous | 83 (96.5) | 289 (98.3) |
| Stage | | |
| II | 7 (8.1) | 26 (8.8) |
| III | 69 (80.2) | 233 (79.3) |
| IV | 10 (11.6) | 35 (11.9) |
| Surgery type | | |
| IDS | 50 (58.1) | 138 (46.9) |
| PDS | 36 (41.9) | 156 (53.1) |
| Surgery residual[a] | | |
| R0 | 50 (58.1) | 183 (62.2) |
| R1 | 28 (32.6) | 79 (26.9) |
| R2 | 8 (9.3) | 32 (10.9) |
| Concurrent use of bevacizumab[b] | | |
| Without | 80 (93) | 271 (92.2) |
| With | 6 (7) | 23 (7.8) |
| Round of chemotherapy[c] | 8 (6-9) | 8 (6-9) |
| Pre-treatment CA125[d], U/ml | 674.55 (262-1549.5) | 705.95 (248.95-1442.08) |

Note: Data are median (IQR) or n (%).

Abbreviations: FIGO, International Federation of Gynecology and Obstetrics; PDS, primary debulking surgery; IDS, interval debulking surgery.

[a]R0, no residual; R1, residual < 1cm; R2, residual ≥ 1cm.

[b]Being with concurrent use of bevacizumab stands for having at least one dose of bevacizumab administered during first-line treatment.

[c]Round of chemotherapy is the sum of round of first-line neoadjuvant chemotherapy and round of first-line adjuvant chemotherapy.

[d]Pre-treatment CA125 stands for the CA125 measured before any form of first-line treatment including surgery and chemotherapy.

**Table S3. Baseline clinical characteristics in FPMT cohort**

| Characteristic | HRD status negative (N=16) | HRD status positive (N=67) |
|---|---|---|
| Age, years | 56 (50-71) | 52 (46-58) |
| Cancer type | | |
| Fallopian tube cancer | 0 (0) | 1 (1.5) |
| Ovarian cancer | 15 (93.8) | 66 (98.5) |
| Primary peritoneal cancer | 1 (6.2) | 0 (0) |
| Histological type | | |
| Grade-3 endometroid | 1 (6.2) | 0 (0) |
| High-grade serous | 15 (93.8) | 67 (100) |
| Stage | | |
| II | 1 (6.2) | 5 (7.5) |
| III | 10 (62.5) | 48 (71.6) |
| IV | 5 (31.2) | 14 (20.9) |
| Surgery type | | |
| IDS | 11 (68.8) | 36 (53.7) |
| PDS | 5 (31.2) | 31 (46.3) |
| Surgery residual[a] | | |
| R0 | 10 (62.5) | 52 (77.6) |
| R1 | 3 (18.8) | 8 (11.9) |
| R2 | 3 (18.8) | 7 (10.4) |
| PARPi type | | |
| Niraparib | 5 (55.6) | 9 (20.5) |
| Olaparib | 4 (44.4) | 35 (79.5) |
| Concurrent use of bevacizumab[b] | | |
| Without | 10 (62.5) | 53 (79.1) |
| With | 6 (37.5) | 14 (20.9) |
| Round of chemotherapy[c] | 6 (6-7) | 6 (6-7) |
| Pre-treatment CA125[d], U/ml | 402.73 (164.4-818.88) | 799 (243.74-1657.77) |
| Time to PARPi[e], days | 37 (31-58) | 46 (37-57) |

Note: Data are median (IQR) or n (%).

Abbreviations: FIGO, International Federation of Gynecology and Obstetrics; PDS, primary debulking surgery; IDS, interval debulking surgery.

[a]R0, no residual; R1, residual < 1cm; R2, residual ≥ 1cm.

[b]Being with concurrent use of bevacizumab stands for having at least one dose of bevacizumab administered during first-line treatment.

[c]Round of chemotherapy is the sum of round of first-line neoadjuvant chemotherapy and round of first-line adjuvant chemotherapy.

[d]Pre-treatment CA125 stands for the CA125 measured before any form of first-line treatment including surgery and chemotherapy.

[e]Time to PARPi stands for the time from last dose of first-line adjuvant chemotherapy to first dose of first-line PARPi maintenance therapy.
